# XBB.1.5 mRNA COVID-19 Vaccination and Inpatient or Emergency Department Visits Among Adults Infected with SARS-CoV-2 JN.1 and XBB-Lineage Variants

**DOI:** 10.1101/2024.03.05.24303796

**Authors:** Matthew E. Levy, Vanessa Chilunda, Phillip R. Heaton, Deran McKeen, Jason D. Goldman, Richard E. Davis, Cynthia A. Schandl, William B. Glen, Lisa M. McEwen, Elizabeth T. Cirulli, Dana Wyman, Andrew Dei Rossi, Hang Dai, Magnus Isaksson, Nicole L. Washington, Tracy Basler, Kevin Tsan, Jason Nguyen, Jimmy Ramirez, Efren Sandoval, William Lee, James Lu, Shishi Luo

## Abstract

Within a multi-state viral genomic surveillance program, we conducted a case-control analysis comparing prior receipt of XBB.1.5-adapted mRNA vaccination between SARS-CoV-2-infected adults with inpatient/ED visits (proxy for severe illness) vs outpatient visits. Among 6,551 patients from September 2023-January 2024, 6.1% with inpatient/ED visits vs 12.0% with outpatient visits had received XBB.1.5 vaccination (aOR=0.41; 95%CI:0.32-0.53). This protective association was weaker among JN.1 (aOR=0.62; 95%CI:0.40-0.96) vs XBB-lineage (aOR=0.28; 95%CI:0.18-0.43) variant infections (interaction, p=0.003). XBB.1.5 vaccination was also protective specifically compared to BA.4/BA.5-adapted mRNA vaccination (aOR=0.60; 95%CI:0.45-0.79). XBB.1.5 vaccines protect against severe illness, but protection may be weaker against JN.1 vs XBB-lineage variants.

## INTRODUCTION

On 11 September 2023, the US Food and Drug Administration (FDA) approved the 2023-2024 BNT162b2 (Pfizer-BioNTech) and mRNA-1273 (Moderna) monovalent mRNA COVID-19 vaccines for individuals aged ≥12 years, with emergency use authorization granted for children aged 6 months to 11 years. These vaccines target the spike protein of the SARS-CoV-2 Omicron variant XBB.1.5, which was predominant in the US from January to May 2023 [1]. The US Centers for Disease Control’s (CDC’s) Advisory Committee on Immunization Practices (ACIP) subsequently recommended that individuals aged ≥6 months receive an XBB.1.5-adapted vaccine regardless of their vaccination history, to enhance protection against circulating variants [2].

By September 2023, other XBB variants such as EG.5 and HV.1 (both sublineages of XBB.1.9.2) had surpassed XBB.1.5 in prevalence. By late December 2023, another novel variant, JN.1 (a sublineage of BA.2.86), had become predominant, accounting for 65% of SARS-CoV-2 infections nationwide by 6 January 2024 [1]. The rapid rise of JN.1, which possesses more than 30 mutations in the spike protein compared to XBB.1.5 (including the notable L455S mutation), could be attributed to increased immune escape and infectivity [3,4]. In laboratory-based neutralization studies, JN.1 has displayed increased resistance to neutralization by antibodies induced by XBB.1.5-adapted mRNA vaccination [4–6]. However, immunogenicity studies suggest that antibody titers are likely to remain effective [7]. Initial findings indicate short-term effectiveness of XBB.1.5-adapted mRNA vaccines for protecting against symptomatic SARS-CoV-2 infection and severe COVID-19 outcomes, largely during periods of XBB predominance [8–14].

To our knowledge, no prior study has examined variant-specific estimates of XBB.1.5 vaccine protection against severe COVID-19 outcomes, and it remains unknown whether XBB.1.5 vaccines protect against severe illness from JN.1 variant infection. Here, we leverage a multi-state viral genomic surveillance program to conduct a case-control analysis assessing the effect of XBB.1.5-adapted mRNA vaccination on patients’ likelihood of having inpatient or emergency department (ED) visits (considered severe) vs outpatient visits (considered mild), among adults with medically attended SARS-CoV-2 infection. Serving as an indicator of vaccine-associated protection against severe illness post-infection, this association was evaluated overall and separately among JN.1 and XBB-lineage variant infections.

## METHODS

### Design and Setting

Within a pan-respiratory virus genomic surveillance program, residual clinical samples from patients who tested positive for a respiratory virus (molecular or antigen) were obtained from three health systems with facilities spanning California, Minnesota, South Carolina, Washington, and Wisconsin (Supplementary Table 1). Samples were initially collected from patients during medical visits and were characterized based on location of collection as inpatient, ED, or outpatient. Patients’ demographic characteristics and COVID-19 vaccination history were extracted from electronic health records (EHRs). Study protocols were approved by institutional review boards.

### Viral Sequencing

Viral sequencing was performed by Helix using a hybridization-capture based assay (Twist Biosciences) and short-read genome sequencing technology (Illumina), as previously described [15]. SARS-CoV-2 was identified in samples with reads that aligned to the reference genome, and lineages were assigned using pangolin version 4.3.1. Further details are provided in the Supplementary Methods.

### Study Sample

This analysis included SARS-CoV-2-positive samples collected from adults aged ≥18 years between 24 September 2023 and 21 January 2024. SARS-CoV-2 infection was identified from either clinical diagnostic testing (performed/ordered by the health system), viral sequencing (performed by Helix), or both.

### Visit Type

The clinical visit type associated with sample collection was a surrogate measure of the severity of illness at time of testing. Inpatient and ED visits represented more severe illness compared to outpatient visits. The reason for the visit and patients’ specific symptoms were not available for analysis.

### Vaccination Status

COVID-19 vaccination status was assigned using the date and type of the most recent dose received prior to the specimen collection date (regardless of the total number of earlier doses received). Patients were considered to be: 1) vaccinated with an XBB.1.5-adapted monovalent mRNA vaccine if their last dose occurred on/after September 12, 2023 and was BNT162b2 or mRNA-1273; 2) vaccinated with a BA.4/BA.5-adapted bivalent mRNA vaccine if their last dose occurred between September 1, 2022 and September 11, 2023 and was BNT162b2 or mRNA-1273; 3) vaccinated with an original wild-type monovalent mRNA or viral vector vaccine if their last dose occurred before September 1, 2022 and was either BNT162b2, mRNA-1273, or Ad26.COV2.S; and 4) unvaccinated if they had received no prior COVID-19 vaccine doses. Patients were excluded if they received any dose 0-6 days before the collection date (83) or if their most recent dose was a different (20) or unknown (124) vaccine type.

### Statistical Analysis

In this case-control analysis, the odds of prior XBB.1.5 vaccination were compared between inpatient or ED patients (cases) and outpatients (controls). We also compared inpatient vs outpatient (excluding ED). Adjusted odds ratios (aORs) and 95% confidence intervals (CIs) for the association between vaccination status and visit type were calculated using multivariable logistic regression, adjusting for age group, sex, race/ethnicity, health system and state of residence, and collection date (natural cubic spline). In multivariable models, effect modification by variant (JN.1 vs XBB-lineage) was assessed using an interaction term with vaccination status. Receipt of an XBB.1.5 vaccine was compared to no receipt of an XBB.1.5 vaccine (irrespective of vaccination history) and to three specific reference groups: 1) BA.4/BA.5 vaccination but no XBB.1.5 vaccine; 2) wild-type vaccination but no BA.4/BA.5 or XBB.1.5 vaccine; and 3) unvaccinated. In a separate analysis, XBB.1.5 vaccine recipients were further categorized based on duration of time since their dose (7-59 vs ≥60 days earlier). In addition, BA.4/BA.5 and wild-type vaccine recipients were compared to unvaccinated. Subgroup analyses were conducted among patients infected with JN.1, XBB-lineage (any), HV.1, and EG.5 variants; among patients aged ≥65 years; and among patients with no other respiratory virus coinfection. *P*<0.05 was considered statistically significant and analyses were performed using R version 4.2.3.

## RESULTS

Among 6,551 adults with medically attended laboratory-confirmed SARS-CoV-2 infection, 1,912 (29.2%) were tested in either an inpatient (1,012; 15.4%) or ED (900; 13.7%) setting, while 4,639 (70.8%) were tested in an outpatient setting. Most SARS-CoV-2 infections were detected through clinical diagnostic testing, with only 8 first identified through viral sequencing. Patient characteristics stratified by visit type are presented in Table 1. Inpatients had a higher median age (73 years; IQR:61-82) compared to ED patients (55 years; IQR:35-72) and outpatients (52 years; IQR:36-68). Lineages were successfully assigned to 4,480 samples (68.4%), with the most prevalent variants being JN.1 (1,084; 24.2%), HV.1 (803; 17.9%), and EG.5 (746; 16.7%). During the most recent 2-week period ending 20 January 2024, JN.1 accounted for 73.1% of sequenced samples, while HV.1 (5.5%) and EG.5 (2.9%) were less prevalent, consistent with national data [1].

**Table 1.** Patient Characteristics Overall and Stratified by Visit Type.

| Characteristic | Visit type |  |  |  |
| --- | --- | --- | --- | --- |
|  | Overall | Inpatient | ED | Outpatient |
|  | (n=6,551),<br>No. (%) | (n=1,012),<br>No. (%) | (n=900),<br>No. (%) | (n=4,639),<br>No. (%) |
| Health system |  |  |  |  |
| HealthPartners | 5,466 (83.4) | 448 (44.3) | 566 (62.9) | 4,452 (96.0) |
| Medical University of South Carolina | 202 (3.1) | 113 (11.2) | 61 (6.8) | 28 (0.6) |
| Providence Health and Services | 883 (13.5) | 451 (44.6) | 273 (30.3) | 159 (3.4) |
| Month of specimen collection |  |  |  |  |
| September 2023 | 410 (6.3) | 62 (6.1) | 62 (6.9) | 286 (6.2) |
| October 2023 | 1,385 (21.1) | 251 (24.8) | 185 (20.6) | 949 (20.5) |
| November 2023 | 1,604 (24.5) | 220 (21.7) | 223 (24.8) | 1,161 (25.0) |
| December 2023 | 2,316 (35.4) | 316 (31.2) | 307 (34.1) | 1,693 (36.5) |
| January 2024 | 836 (12.8) | 163 (16.1) | 123 (13.7) | 550 (11.9) |
| Age group |  |  |  |  |
| 18 to 49 y | 2,684 (41.0) | 138 (13.6) | 387 (43.0) | 2,159 (46.5) |
| 50 to 64 y | 1,428 (21.8) | 163 (16.1) | 176 (19.6) | 1,089 (23.5) |
| 65 to 74 y | 1,106 (16.9) | 246 (24.3) | 137 (15.2) | 723 (15.6) |
| 75 to 84 y | 904 (13.8) | 270 (26.7) | 135 (15.0) | 499 (10.8) |
| ≥85 y | 429 (6.5) | 195 (19.3) | 65 (7.2) | 169 (3.6) |
| Female | 3,893 (59.4) | 505 (49.9) | 523 (58.1) | 2,865 (61.8) |
| Race and ethnicity |  |  |  |  |
| Asian, non-Hispanic | 325 (5.0) | 23 (2.3) | 20 (2.2) | 282 (6.1) |
| Black, non-Hispanic | 828 (12.6) | 88 (8.7) | 146 (16.2) | 594 (12.8) |
| Hispanic | 256 (3.9) | 18 (1.8) | 41 (4.6) | 197 (4.2) |
| White, non-Hispanic | 3,851 (58.8) | 432 (42.7) | 413 (45.9) | 3,006 (64.8) |
| Other or unknown | 1,291 (19.7) | 451 (44.6) | 280 (31.1) | 560 (12.1) |
| COVID-19 vaccination status<sup>a</sup> |  |  |  |  |
| Unvaccinated | 1,842 (28.1) | 360 (35.6) | 358 (39.8) | 1,124 (24.2) |
| Vaccinated with a wild-type vaccine | 2,879 (43.9) | 397 (39.2) | 386 (42.9) | 2,096 (45.2) |
| Vaccinated with a BA.4/BA.5 vaccine | 1,155 (17.6) | 182 (18.0) | 112 (12.4) | 861 (18.6) |
| Vaccinated with an XBB.1.5 vaccine | 675 (10.3) | 73 (7.2) | 44 (4.9) | 558 (12.0) |
| SARS-CoV-2 variant<sup>b</sup> |  |  |  |  |
| JN.1 | 1,084 (16.5) | 139 (13.7) | 165 (18.3) | 780 (16.8) |
| HV.1 | 803 (12.3) | 116 (11.5) | 114 (12.7) | 573 (12.4) |
| EG.5 | 746 (11.4) | 119 (11.8) | 119 (13.2) | 508 (11.0) |
| XBB.1.16.1 | 379 (5.8) | 54 (5.3) | 67 (7.4) | 258 (5.6) |
| XBB.1.5 | 262 (4.0) | 44 (4.3) | 47 (5.2) | 171 (3.7) |
| HK.3 | 231 (3.5) | 39 (3.9) | 33 (3.7) | 159 (3.4) |
| XBB.1.16.6 | 219 (3.3) | 34 (3.4) | 43 (4.8) | 142 (3.1) |
| XBB.2.3 | 209 (3.2) | 31 (3.1) | 24 (2.7) | 154 (3.3) |
| FL.1.5.1 | 151 (2.3) | 24 (2.4) | 26 (2.9) | 101 (2.2) |
| BA.2.86 | 146 (2.2) | 20 (2.0) | 23 (2.6) | 103 (2.2) |
| XBB.1.9.1 | 38 (0.6) | 6 (0.6) | 5 (0.6) | 27 (0.6) |
| XBB.1.16 | 37 (0.6) | 5 (0.5) | 7 (0.8) | 25 (0.5) |
| XBB.1.9.2 | 20 (0.3) | 7 (0.7) | 3 (0.3) | 10 (0.2) |
| Other XBB | 98 (1.5) | 15 (1.5) | 9 (1.0) | 74 (1.6) |
| Other non-XBB | 57 (0.9) | 6 (0.6) | 9 (1.0) | 42 (0.9) |
| Insufficient sequencing data | 2,071 (31.6) | 353 (34.9) | 206 (22.9) | 1,512 (32.6) |

Abbreviations: ED, emergency department.
<sup>a</sup> Defined by whether there was $\geq 1$ COVID-19 vaccine record prior to the specimen collection date as well as by the date and type of the most recent dose received.
<sup>b</sup> SARS-CoV-2 variant lineages were assigned using pangolin version 4.3.1. Except for HV.1 and HK.3, sublineages of EG.5 are aggregated with EG.5. Except for JN.1, sublineages of BA.2.86 are aggregated with BA.2.86. Except for FL.1.5.1, sublineages of XBB.1.9.1 are aggregated with XBB.1.9.1. Except for XBB.1.16.1 and XBB.1.16.6, sublineages of XBB.1.16 are aggregated with XBB.1.16. Except for EG.5, HV.1, and HK.3, sublineages of XBB.1.9.2 are aggregated with XBB.1.9.2. Sublineages of each other named lineage are aggregated with the respective lineage. JN.1 is also known as BA.2.86.1.1; HV.1 as XBB.1.9.2.5.1.6.1; EG.5 as XBB.1.9.2.5; HK.3 as XBB.1.9.2.5.1.1.3; and FL.1.5.1 as XBB.1.9.1.1.5.1.

Regarding vaccination status, 675 (10.3%) patients had received XBB.1.5 vaccination (a median of 57 days earlier [IQR:39-73; range:7-122]), 1,155 (17.6%) had received BA.4/BA.5 vaccination (but not XBB.1.5 vaccination) (median of 374 days since last dose [IQR:330-414]), 2,879 (43.9%) had received wild-type vaccination (but not BA.4/BA.5 or XBB.1.5 vaccination) (median of 712 days since last dose [IQR:625-818]), and 1,842 (28.1%) were unvaccinated. Among XBB.1.5-vaccinated patients, the median time since vaccination was 64 days for JN.1 infections (IQR:51-80) and 52 days for XBB-lineage infections (IQR:31-64). In the most recent 14-day period, 19.4% of patients overall were XBB.1.5-vaccinated.

Among all SARS-CoV-2 infections, 6.1% of patients with inpatient/ED visits had received XBB.1.5 vaccination, compared to 12.0% of patients with outpatient visits (Figure 1). In multivariable analysis, XBB.1.5 vaccination vs no XBB.1.5 vaccination was associated with lower odds of having inpatient/ED visits compared to outpatient visits (aOR=0.41; 95%CI:0.32-0.53). This protective association was significant among any-variant infections regardless of the specific reference group used: vs BA.4/BA.5 vaccination (aOR=0.60; 95%CI:0.45-0.79); vs wild-type vaccination (aOR=0.48; 95%CI:0.37-0.63); and vs unvaccinated (aOR=0.24; 95%CI:0.19-0.32).

**Figure 1.**
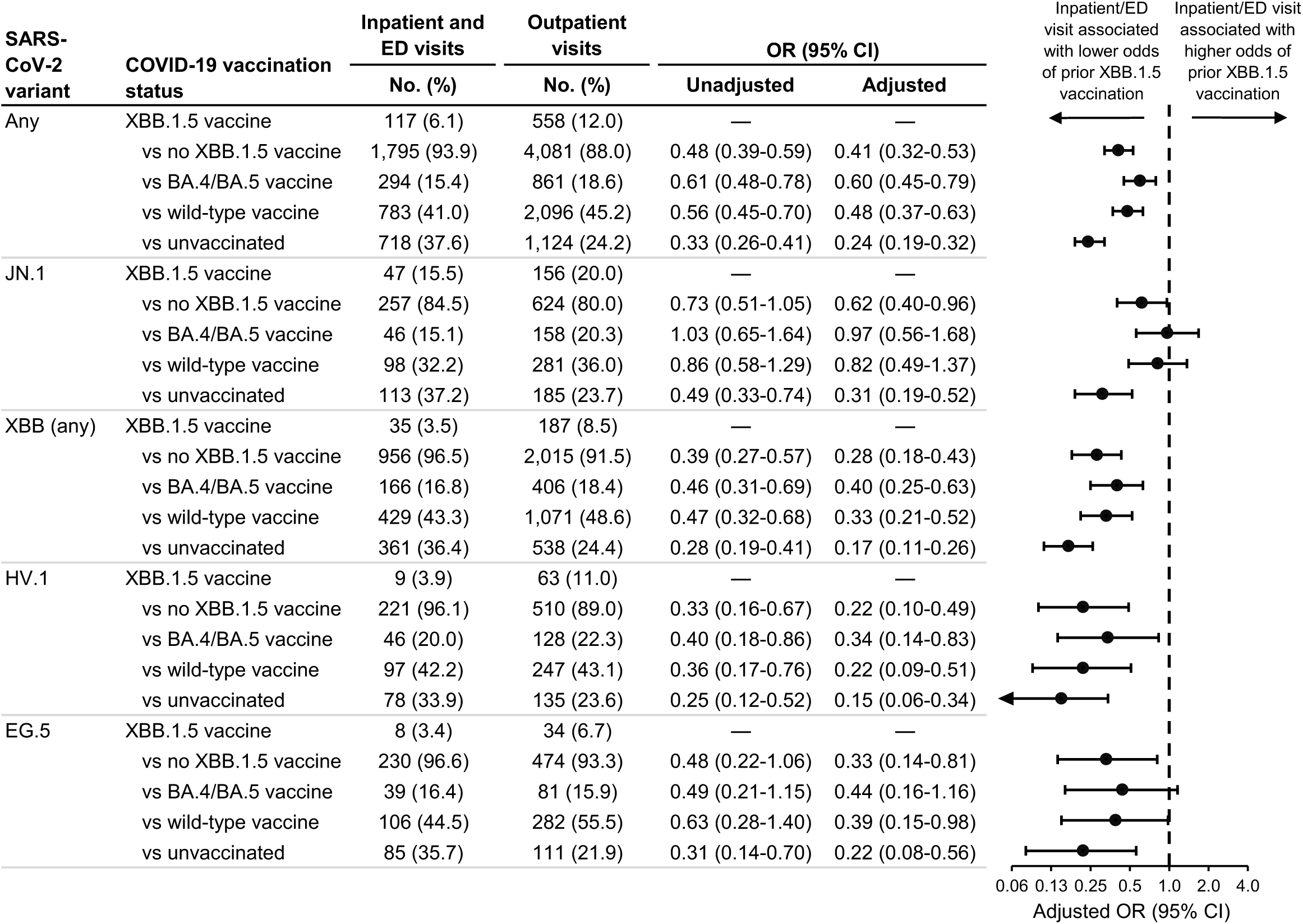
Association between inpatient or emergency department vs outpatient visit type and prior receipt of an XBB.1.5 vaccine. Associations were calculated among all SARS-CoV-2 infections and among JN.1, XBB (any sublineage), HV.1, and EG.5 infections. Odds ratios (ORs) were calculated comparing prior receipt of an XBB.1.5 vaccine to no prior receipt of an XBB.1.5 vaccine (irrespective of previous COVID-19 vaccination history) as well as to each of three specific reference groups: 1) prior receipt of a BA.4/BA.5 vaccine but not an XBB.1.5 vaccine; 2) prior receipt of a wild-type vaccine but not a BA.4/BA.5 or XBB.1.5 vaccine; and 3) unvaccinated.. Adjusted ORs were adjusted for age group (18-49, 50-64, 65-74, 75-84, and ≥85 years), sex, race and ethnicity (Asian, non-Hispanic; Black, non-Hispanic; Hispanic; white, non-Hispanic; and other/unknown), health system and state of residence, and collection date (natural cubic spline with 4 degrees of freedom). CI indicates confidence interval; ED, emergency department.

When stratifying by variant, patients with XBB.1.5 vaccination (vs. no XBB.1.5 vaccination) had lower odds of inpatient/ED visits among JN.1 infections (aOR=0.62; 95%CI:0.40-0.96) and among XBB-lineage infections (aOR=0.28; 95%CI:0.18-0.43) (Figure 1), but this association was weaker among JN.1 vs XBB-lineage infections (vaccination-variant interaction, p=0.003). This interaction between vaccination and JN.1 vs XBB-lineage variant remained significant when comparing XBB.1.5-vaccinated patients to BA.4/BA.5-vaccinated (p=0.009), wild-type-vaccinated (p=0.003), and unvaccinated (p=0.035) patients. Regarding specific vaccination status reference groups, protective associations for XBB.1.5 vaccination were strongest when comparing XBB.1.5-vaccinated to unvaccinated patients.

In additional analyses, findings were similar when only including adults aged ≥65 years (Supplementary Figure 1; interaction between XBB.1.5 vaccination and JN.1 vs XBB-lineage variant, p=0.043), when comparing only inpatient vs outpatient visits (Supplementary Figure 2; interaction, p=0.013), and when excluding patients with respiratory virus coinfections (Supplementary Figure 3; interaction, p=0.005). Among JN.1 and XBB-lineage infections, similar protective associations were detected for patients XBB.1.5-vaccinated 7-59 days earlier and ≥60 days earlier (Supplementary Figure 4). BA.4/BA.5 vaccination and wild-type vaccination were also associated with lower odds of inpatient/ED visits compared to unvaccinated (Supplementary Figure 5).

## DISCUSSION

In this multi-state study of adults with medically attended SARS-CoV-2 infection between September 2023 and January 2024, XBB.1.5 mRNA-vaccinated individuals had an overall 59% lower odds of having an inpatient/ED visit vs outpatient visit. Unlike previous observational studies examining XBB.1.5 vaccination and severe COVID-19 outcomes [8,9,11,14], our study provides novel data on XBB.1.5 vaccine-associated protection against specific contemporary circulating variants, which was made possible through the linkage of viral sequencing and patient-level EHR data. While findings provide evidence that XBB.1.5 vaccines protected against severe illness associated with both JN.1 and XBB-lineage variants, protection against JN.1 was significantly lower. Among JN.1-infected patients, XBB.1.5 vaccination was associated with 38% lower odds of inpatient/ED visits, compared to the 72% lower odds observed among XBB-lineage-infected patients. Given that associations among JN.1-infected individuals were similar irrespective of time since XBB.1.5 vaccination, this difference by variant was not attributed to waning effectiveness over the approximately 4 months of available data. Similarly, a study evaluating XBB.1.5 vaccine effectiveness against symptomatic infection reported lower point estimates for likely-JN.1 infections (non-confirmed) than for non-JN.1 lineages [10]. This key finding emphasizes the need for COVID-19 vaccines to be routinely updated to align with circulating strains and for individuals to stay up to date with recommended vaccines.

The inverse association between XBB.1.5 vaccination (administered a median of 57 days earlier) and inpatient/ED visit type was consistently observed regardless of the specific reference group used, which included prior receipt of BA.4/BA.5 vaccination without XBB.1.5 vaccination. Thus, XBB.1.5-vaccine-associated protection was enhanced beyond the remaining immunity conferred by previous BA.4/BA.5 vaccination. It is important to note that results were obtained within the context of widespread natural immunity from prior infection, which could have resulted in attenuated associations if individuals who had not received XBB.1.5 vaccination were more likely to have infection-induced immunity.

This study has several limitations. First, although all patients had laboratory-confirmed SARS-CoV-2 infection identified at a medical visit, we were unable to confirm symptomatic COVID-19 and we lacked data on the reasons for medical visits. Second, visit type was solely assessed at the time of sample collection for SARS-CoV-2 testing and it is unknown whether outpatients subsequently had ED or inpatient visits later in the course of infection, potentially contributing to misclassification. Third, we lacked data on social factors (e.g., insurance type) and underlying medical conditions, which could have resulted in residual confounding. Fourth, misclassification of vaccination status is possible if vaccine doses documented in EHRs were not complete. Finally, because vaccination status subgroups were distinct both in vaccine type received and in time from last vaccination, it is not possible to discern whether the protective effect shown here is attributable primarily to vaccine type or titers of neutralizing antibodies.

The findings from this study provide evidence of the effectiveness of the XBB.1.5-adapted mRNA vaccines in protecting against severe illness requiring inpatient or ED visits among adults infected with JN.1 or XBB-lineage variants. These results support the current recommendation that all adults should, irrespective of their previous COVID-19 vaccination history, receive the 2023-2024 COVID-19 vaccine to enhance their protection. However, findings also suggest that XBB.1.5 vaccines provide comparatively less protection against the currently predominant JN.1 variant than they do against XBB-lineage variants. Future research is needed to confirm the degree of XBB.1.5 vaccine protection against severe illness associated with JN.1 variant infection and to evaluate the potential for waning effectiveness over time.

## NOTES

### Author contributions

All authors contributed substantively to this manuscript in the following ways: conceptualization (MEL, VC, SL), investigation (MEL, VC, PRH, DM, JDG, RED, CAS, WBG, LMM, DW, ADR, HD, MI, NLW, TB, KT, JN, JR, ES, SL), data curation (MEL, VC, LMM, DW, ADR, HD, MI, NLW, SL), formal analysis (MEL), project administration (PRH, JDG, CAS, ETC, NLW, WL, JL, SL), writing - original draft (MEL), and writing - review and editing (MEL, VC, PRH, DM, JDG, RED, CAS, WBG, ETC, SL).

## Data Availability

Data collected for this study are not available. Data sharing agreements between Helix and partner institutions prohibit Helix from making this dataset publicly available.

## Acknowledgements

The authors thank the Helix Clinical Informatics and Bioinformatics teams for their contributions to electronic health record data and viral sequencing pipelines. They also thank Catherine Clinton for her oversight and guidance in ensuring research compliance. They thank the investigators and staff at HealthPartners, Providence Health, and the Medical University of South Carolina who contributed to the ViEW Network^TM^.

