## Supplement for "XBB.1.5 mRNA COVID-19 Vaccination and Inpatient or Emergency Department Visits Among Adults Infected with SARS-CoV-2 JN.1 and XBB-Lineage Variants"

**Supplementary Methods:**

**Viral sequencing, bioinformatic analyses, and SARS-CoV-2 lineage designation**

### Viral sequencing

RNA was extracted from 400 μL of remnant patient anterior nares samples using the MagMAX Viral/Pathogen kit (ThermoScientific). RNA libraries (5μL RNA input volume) were generated using the Rapid RNA Library Kit protocol (Swift Biosciences/Integrated DNA Technologies). SARS-CoV-2 genome was enriched and captured using the Respiratory Virus Research Panel hybridization probe panel (Twist Biosciences). Samples were sequenced using the NovaSeq 6000 Sequencing system S1 flow cell, with S1 Reagent Kit v1.5 (300 cycles).

### Bioinformatic analyses

The flow cell output was demultiplexed with bcl2fastq (Illumina) into per-sample FASTQ sequences. These sequences were then processed using the Helix fastagenerator pipeline to produce a consensus sequence FASTA file. First, reads were aligned to a reference set consisting of a representative genome of each respiratory virus that was targeted by the hybridization probes, and the human transcriptome (GENCODE v37) using BWA-MEM. The non-human reads were then re-aligned to an expanded set of respiratory viruses reference genomes including the SARS-CoV-2 genome (NCBI accession NC_045512.2) using a de Bruijn graph based algorithm (based on https://doi.org/10.1093/bioinformatics/btz814) to identify sequences that best match the reads. SARS-CoV-2 aligned reads were then marked for duplicates followed by variant calling using the Haplotyper algorithm (Sentieon, Inc). The consensus sequence for each sample was generated based on per-base coverage from the alignment file (BAM) and per-variant allele depths from the variant call format (VCF) file according to the following criteria: coverage from at least 5 unique reads with at least 80% of the reads supporting the allele. For the bases that did not meet this criteria, an N was reported. A sequence is considered to pass quality criteria if it contains at most 30% N bases.

**SARS-CoV-2 lineage designation**

Viral sequences were assigned a PANGO lineage using pangolin (https://github.com/cov-lineages/pangolin). For this analysis, Pangolin data version v1.23.1 with Pangolin software v4.3.1 was used.

**Supplementary Table 1. Health Systems Participating in the ViEW Network™.**

| Health system | **HealthPartners** | **Providence Health** | **Medical University of South Carolina (MUSC)** |
|---|---|---|---|
| Protocol number | 0006-001 | 0006-001 | 0006-001 |
| Institutional Review Board (IRB) | WCG IRB | WCG IRB | MUSC Institutional Review Board for Human Research |
| IRB approval number | 20224919 | 20224919 | Pro00129083 |
| Geographic region | Minnesota and western Wisconsin | Eastern Washington and southern California | South Carolina |
| Sample size | 5,466 | 883 | 202 |
| Date range of patients included | September 24, 2023–January 20, 2024 | September 24, 2023–January 21, 2024 | September 24, 2023–January 13, 2024 |
| Informed consent | Waiver of consent obtained | Waiver of consent obtained | Waiver of consent obtained |
| Data privacy | Limited dataset stripped of direct identifiers | Limited dataset stripped of direct identifiers | Limited dataset stripped of direct identifiers |

| SARS-CoV-2 variant | COVID-19 vaccination status | Inpatient and ED visits | Outpatient visits | OR (95% CI) | |
|---|---|---|---|---|---|
| | | No. (%) | No. (%) | Unadjusted | Adjusted |
| Any | XBB.1.5 vaccine | 96 (9.2) | 310 (22.3) | — | — |
| | vs no XBB.1.5 vaccine | 952 (90.8) | 1,081 (77.7) | 0.35 (0.28-0.45) | 0.39 (0.29-0.52) |
| | vs BA.4/BA.5 vaccine | 220 (21.0) | 323 (23.2) | 0.45 (0.34-0.61) | 0.50 (0.36-0.70) |
| | vs wild-type vaccine | 423 (40.4) | 527 (37.9) | 0.39 (0.30-0.50) | 0.44 (0.32-0.60) |
| | vs unvaccinated | 309 (29.5) | 231 (16.6) | 0.23 (0.17-0.31) | 0.24 (0.17-0.34) |
| JN.1 | XBB.1.5 vaccine | 40 (23.0) | 94 (39.8) | — | — |
| | vs no XBB.1.5 vaccine | 134 (77.0) | 142 (60.2) | 0.45 (0.29-0.70) | 0.52 (0.31-0.86) |
| | vs BA.4/BA.5 vaccine | 33 (19.0) | 58 (24.6) | 0.75 (0.42-1.32) | 0.84 (0.44-1.63) |
| | vs wild-type vaccine | 45 (25.9) | 54 (22.9) | 0.51 (0.30-0.88) | 0.68 (0.35-1.30) |
| | vs unvaccinated | 56 (32.2) | 30 (12.7) | 0.23 (0.13-0.41) | 0.20 (0.10-0.41) |
| XBB (any) | XBB.1.5 vaccine | 28 (4.9) | 117 (15.9) | — | — |
| | vs no XBB.1.5 vaccine | 540 (95.1) | 618 (84.1) | 0.27 (0.18-0.42) | 0.26 (0.16-0.42) |
| | vs BA.4/BA.5 vaccine | 134 (23.6) | 175 (23.8) | 0.31 (0.20-0.50) | 0.33 (0.20-0.56) |
| | vs wild-type vaccine | 246 (43.3) | 303 (41.2) | 0.29 (0.19-0.46) | 0.29 (0.17-0.48) |
| | vs unvaccinated | 160 (28.2) | 140 (19.0) | 0.21 (0.13-0.34) | 0.18 (0.10-0.30) |
| HV.1 | XBB.1.5 vaccine | 7 (5.0) | 40 (20.6) | — | — |
| | vs no XBB.1.5 vaccine | 134 (95.0) | 154 (79.4) | 0.20 (0.09-0.46) | 0.14 (0.06-0.38) |
| | vs BA.4/BA.5 vaccine | 38 (27.0) | 49 (25.3) | 0.23 (0.09-0.56) | 0.18 (0.06-0.50) |
| | vs wild-type vaccine | 55 (39.0) | 62 (32.0) | 0.20 (0.08-0.48) | 0.13 (0.05-0.38) |
| | vs unvaccinated | 41 (29.1) | 43 (22.2) | 0.18 (0.07-0.46) | 0.13 (0.04-0.36) |
| EG.5 | XBB.1.5 vaccine | 8 (6.2) | 21 (11.9) | — | — |
| | vs no XBB.1.5 vaccine | 121 (93.8) | 156 (88.1) | 0.49 (0.21-1.15) | 0.34 (0.12-0.96) |
| | vs BA.4/BA.5 vaccine | 32 (24.8) | 38 (21.5) | 0.45 (0.18-1.16) | 0.47 (0.15-1.45) |
| | vs wild-type vaccine | 51 (39.5) | 92 (52.0) | 0.69 (0.28-1.66) | 0.45 (0.15-1.35) |
| | vs unvaccinated | 38 (29.5) | 26 (14.7) | 0.26 (0.10-0.68) | 0.16 (0.05-0.52) |

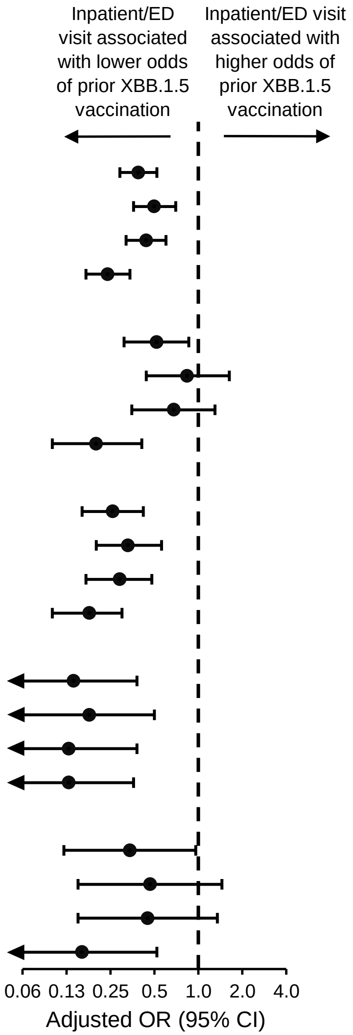


**Supplementary Figure 1. Association between inpatient or emergency department vs outpatient visit type and prior receipt of an XBB.1.5 vaccine among older adults aged ≥65 years.** Associations were calculated among all SARS-CoV-2 infections and among JN.1, XBB (any sublineage), HV.1, and EG.5 infections. Odds ratios (ORs) were calculated comparing prior receipt of an XBB.1.5 vaccine to no prior receipt of an XBB.1.5 vaccine (irrespective of previous COVID-19 vaccination history) as well as to each of three specific reference groups: 1) prior receipt of a BA.4/BA.5 vaccine but not an XBB.1.5 vaccine; 2) prior receipt of a wild-type vaccine but not a BA.4/BA.5 or XBB.1.5 vaccine; and 3) unvaccinated. Adjusted ORs were adjusted for age group (18-49, 50-64, 65-74, 75-84, and ≥85 years), sex, race and ethnicity (Asian, non-Hispanic; Black, non-Hispanic; Hispanic; white, non-Hispanic; and other/unknown), health system and state of residence, and collection date (natural cubic spline with 4 degrees of freedom). CI indicates confidence interval; ED, emergency department.

| SARS-CoV-2 variant | COVID-19 vaccination status | Inpatient visits | Outpatient visits | OR (95% CI) | |
|---|---|---|---|---|---|
| | | No. (%) | No. (%) | Unadjusted | Adjusted |
| Any | XBB.1.5 vaccine | 73 (7.2) | 558 (12.0) | — | — |
| | vs no XBB.1.5 vaccine | 939 (92.8) | 4,081 (88.0) | 0.57 (0.44-0.73) | 0.48 (0.36-0.66) |
| | vs BA.4/BA.5 vaccine | 182 (18.0) | 861 (18.6) | 0.62 (0.46-0.83) | 0.62 (0.44-0.89) |
| | vs wild-type vaccine | 397 (39.2) | 2,096 (45.2) | 0.69 (0.53-0.90) | 0.60 (0.43-0.84) |
| | vs unvaccinated | 360 (35.6) | 1,124 (24.2) | 0.41 (0.31-0.54) | 0.29 (0.21-0.41) |
| JN.1 | XBB.1.5 vaccine | 27 (19.4) | 156 (20.0) | — | — |
| | vs no XBB.1.5 vaccine | 112 (80.6) | 624 (80.0) | 0.96 (0.61-1.52) | 0.71 (0.40-1.24) |
| | vs BA.4/BA.5 vaccine | 26 (18.7) | 158 (20.3) | 1.05 (0.59-1.88) | 0.89 (0.44-1.81) |
| | vs wild-type vaccine | 34 (24.5) | 281 (36.0) | 1.43 (0.83-2.46) | 1.47 (0.69-3.13) |
| | vs unvaccinated | 52 (37.4) | 185 (23.7) | 0.62 (0.37-1.03) | 0.31 (0.16-0.60) |
| XBB (any) | XBB.1.5 vaccine | 23 (4.7) | 187 (8.5) | — | — |
| | vs no XBB.1.5 vaccine | 471 (95.3) | 2,015 (91.5) | 0.53 (0.34-0.82) | 0.31 (0.19-0.53) |
| | vs BA.4/BA.5 vaccine | 97 (19.6) | 406 (18.4) | 0.51 (0.32-0.84) | 0.42 (0.24-0.75) |
| | vs wild-type vaccine | 211 (42.7) | 1,071 (48.6) | 0.62 (0.40-0.99) | 0.37 (0.21-0.64) |
| | vs unvaccinated | 163 (33.0) | 538 (24.4) | 0.41 (0.25-0.65) | 0.20 (0.11-0.34) |
| HV.1 | XBB.1.5 vaccine | 6 (5.2) | 63 (11.0) | — | — |
| | vs no XBB.1.5 vaccine | 110 (94.8) | 510 (89.0) | 0.44 (0.19-1.05) | 0.25 (0.09-0.67) |
| | vs BA.4/BA.5 vaccine | 25 (21.6) | 128 (22.3) | 0.49 (0.19-1.25) | 0.37 (0.12-1.11) |
| | vs wild-type vaccine | 47 (40.5) | 247 (43.1) | 0.50 (0.20-1.22) | 0.22 (0.08-0.65) |
| | vs unvaccinated | 38 (32.8) | 135 (23.6) | 0.34 (0.14-0.84) | 0.18 (0.06-0.53) |
| EG.5 | XBB.1.5 vaccine | 7 (5.9) | 34 (6.7) | — | — |
| | vs no XBB.1.5 vaccine | 112 (94.1) | 474 (93.3) | 0.87 (0.38-2.02) | 0.38 (0.13-1.11) |
| | vs BA.4/BA.5 vaccine | 24 (20.2) | 81 (15.9) | 0.69 (0.27-1.77) | 0.56 (0.17-1.83) |
| | vs wild-type vaccine | 53 (44.5) | 282 (55.5) | 1.10 (0.46-2.60) | 0.41 (0.13-1.29) |
| | vs unvaccinated | 35 (29.4) | 111 (21.9) | 0.65 (0.27-1.60) | 0.22 (0.07-0.72) |

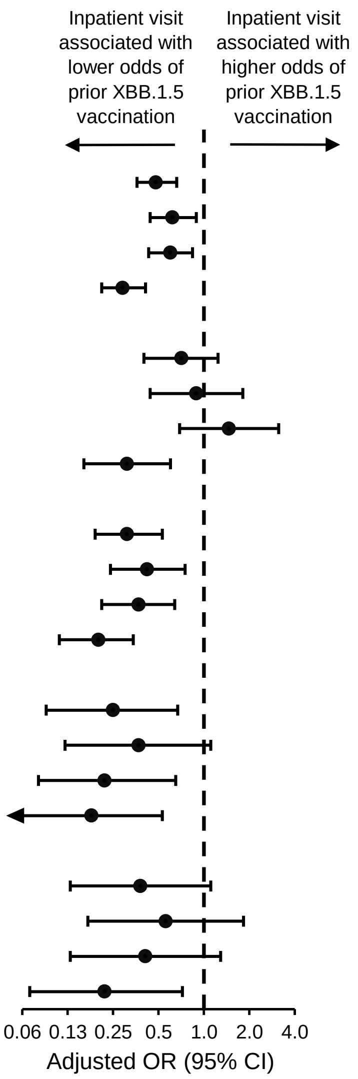


**Supplementary Figure 2. Association between inpatient vs outpatient visit type and prior receipt of an XBB.1.5 vaccine.** Associations were calculated among all SARS-CoV-2 infections and among JN.1, XBB (any sublineage), HV.1, and EG.5 infections. Odds ratios (ORs) were calculated comparing prior receipt of an XBB.1.5 vaccine to no prior receipt of an XBB.1.5 vaccine (irrespective of previous COVID-19 vaccination history) as well as to each of three specific reference groups: 1) prior receipt of a BA.4/BA.5 vaccine but not an XBB.1.5 vaccine; 2) prior receipt of a wild-type vaccine but not a BA.4/BA.5 or XBB.1.5 vaccine; and 3) unvaccinated. Adjusted ORs were adjusted for age group (18-49, 50-64, 65-74, 75-84, and ≥85 years), sex, race and ethnicity (Asian, non-Hispanic; Black, non-Hispanic; Hispanic; white, non-Hispanic; and other/unknown), health system and state of residence, and collection date (natural cubic spline with 4 degrees of freedom). CI indicates confidence interval.

| SARS-CoV-2 variant | COVID-19 vaccination status | Inpatient and ED visits | Outpatient visits | OR (95% CI) | |
|---|---|---|---|---|---|
| | | No. (%) | No. (%) | Unadjusted | Adjusted |
| Any | XBB.1.5 vaccine | 113 (6.1) | 538 (11.9) | — | — |
| | vs no XBB.1.5 vaccine | 1,748 (93.9) | 3,978 (88.1) | 0.48 (0.39-0.59) | 0.42 (0.32-0.54) |
| | vs BA.4/BA.5 vaccine | 292 (15.7) | 847 (18.8) | 0.61 (0.48-0.78) | 0.59 (0.45-0.79) |
| | vs wild-type vaccine | 757 (40.7) | 2,044 (45.3) | 0.57 (0.46-0.71) | 0.49 (0.38-0.64) |
| | vs unvaccinated | 699 (37.6) | 1,087 (24.1) | 0.33 (0.26-0.41) | 0.24 (0.18-0.32) |
| JN.1 | XBB.1.5 vaccine | 47 (15.7) | 154 (20.2) | — | — |
| | vs no XBB.1.5 vaccine | 253 (84.3) | 608 (79.8) | 0.73 (0.51-1.05) | 0.62 (0.40-0.95) |
| | vs BA.4/BA.5 vaccine | 45 (15.0) | 156 (20.5) | 1.06 (0.66-1.69) | 0.98 (0.56-1.69) |
| | vs wild-type vaccine | 97 (32.3) | 273 (35.8) | 0.86 (0.58-1.28) | 0.80 (0.48-1.33) |
| | vs unvaccinated | 111 (37.0) | 179 (23.5) | 0.49 (0.33-0.74) | 0.31 (0.18-0.51) |
| XBB (any) | XBB.1.5 vaccine | 35 (3.6) | 185 (8.5) | — | — |
| | vs no XBB.1.5 vaccine | 943 (96.4) | 1,989 (91.5) | 0.40 (0.28-0.58) | 0.29 (0.19-0.44) |
| | vs BA.4/BA.5 vaccine | 166 (17.0) | 402 (18.5) | 0.46 (0.31-0.69) | 0.40 (0.25-0.64) |
| | vs wild-type vaccine | 423 (43.3) | 1,054 (48.5) | 0.47 (0.32-0.69) | 0.34 (0.22-0.53) |
| | vs unvaccinated | 354 (36.2) | 533 (24.5) | 0.28 (0.19-0.42) | 0.17 (0.11-0.27) |
| HV.1 | XBB.1.5 vaccine | 9 (3.9) | 63 (11.2) | — | — |
| | vs no XBB.1.5 vaccine | 219 (96.1) | 498 (88.8) | 0.32 (0.16-0.66) | 0.22 (0.10-0.49) |
| | vs BA.4/BA.5 vaccine | 46 (20.2) | 127 (22.6) | 0.39 (0.18-0.86) | 0.34 (0.14-0.82) |
| | vs wild-type vaccine | 96 (42.1) | 238 (42.4) | 0.35 (0.17-0.74) | 0.22 (0.09-0.51) |
| | vs unvaccinated | 77 (33.8) | 133 (23.7) | 0.25 (0.12-0.52) | 0.15 (0.06-0.35) |
| EG.5 | XBB.1.5 vaccine | 8 (3.4) | 34 (6.7) | — | — |
| | vs no XBB.1.5 vaccine | 226 (96.6) | 471 (93.3) | 0.49 (0.22-1.08) | 0.34 (0.14-0.83) |
| | vs BA.4/BA.5 vaccine | 39 (16.7) | 81 (16.0) | 0.49 (0.21-1.15) | 0.44 (0.17-1.18) |
| | vs wild-type vaccine | 103 (44.0) | 279 (55.2) | 0.64 (0.29-1.42) | 0.40 (0.16-1.02) |
| | vs unvaccinated | 84 (35.9) | 111 (22.0) | 0.31 (0.14-0.71) | 0.22 (0.08-0.56) |

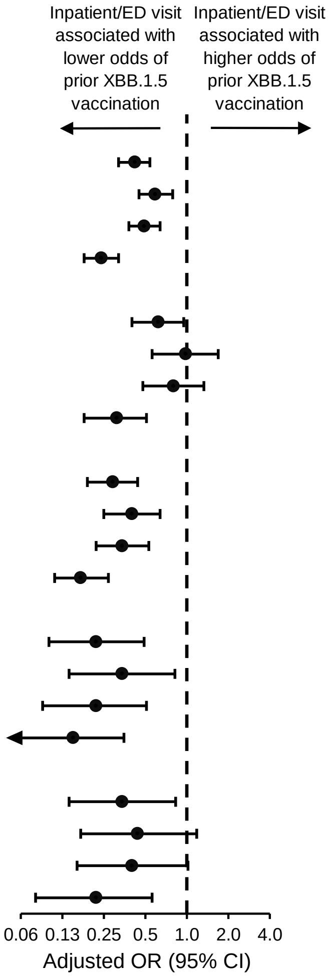


**Supplementary Figure 3. Association between inpatient or emergency department vs outpatient visit type and prior receipt of an XBB.1.5 vaccine among SARS-CoV-2 mono-infected patients.** Patients with a known respiratory virus coinfection were excluded (174), which included influenza (70), rhinovirus/enterovirus (42), RSV (28), human coronavirus (24), parainfluenza (9), adenovirus (3), human metapneumovirus (1), and bocavirus (1) coinfections. Associations were calculated among all SARS-CoV-2 infections and among JN.1, XBB (any sublineage), HV.1, and EG.5 infections. Odds ratios (ORs) were calculated comparing prior receipt of an XBB.1.5 vaccine to no prior receipt of an XBB.1.5 vaccine (irrespective of previous COVID-19 vaccination history) as well as to each of three specific reference groups: 1) prior receipt of a BA.4/BA.5 vaccine but not an XBB.1.5 vaccine; 2) prior receipt of a wild-type vaccine but not a BA.4/BA.5 or XBB.1.5 vaccine; and 3) unvaccinated. Adjusted ORs were adjusted for age group (18-49, 50-64, 65-74, 75-84, and ≥85 years), sex, race and ethnicity (Asian, non-Hispanic; Black, non-Hispanic; Hispanic; white, non-Hispanic; and other/unknown), health system and state of residence, and collection date (natural cubic spline with 4 degrees of freedom). CI indicates confidence interval; ED, emergency department.

| SARS-CoV-2 variant | COVID-19 vaccination status | Inpatient and ED visits | Outpatient visits | OR (95% CI) | |
|---|---|---|---|---|---|
| | | No. (%) | No. (%) | Unadjusted | Adjusted |
| Any | XBB.1.5 vaccine 7-59 days earlier | 54 (2.8) | 309 (6.7) | — | — |
| | vs no XBB.1.5 vaccine | 1,795 (93.9) | 4,081 (88.0) | 0.40 (0.30-0.53) | 0.35 (0.25-0.50) |
| | vs BA.4/BA.5 vaccine | 294 (15.4) | 861 (18.6) | 0.51 (0.37-0.70) | 0.51 (0.35-0.74) |
| | vs wild-type vaccine | 783 (41.0) | 2,096 (45.2) | 0.47 (0.35-0.63) | 0.41 (0.29-0.59) |
| | vs unvaccinated | 718 (37.6) | 1,124 (24.2) | 0.27 (0.20-0.37) | 0.21 (0.14-0.30) |
| | XBB.1.5 vaccine ≥60 days earlier | 63 (3.3) | 249 (5.4) | — | — |
| | vs no XBB.1.5 vaccine | 1,795 (93.9) | 4,081 (88.0) | 0.58 (0.43-0.76) | 0.49 (0.35-0.68) |
| | vs BA.4/BA.5 vaccine | 294 (15.4) | 861 (18.6) | 0.74 (0.55-1.01) | 0.71 (0.49-1.02) |
| | vs wild-type vaccine | 783 (41.0) | 2,096 (45.2) | 0.68 (0.51-0.90) | 0.58 (0.41-0.82) |
| | vs unvaccinated | 718 (37.6) | 1,124 (24.2) | 0.40 (0.30-0.53) | 0.29 (0.20-0.41) |
| JN.1 | XBB.1.5 vaccine 7-59 days earlier | 18 (5.9) | 66 (8.5) | — | — |
| | vs no XBB.1.5 vaccine | 257 (84.5) | 624 (80.0) | 0.66 (0.39-1.14) | 0.65 (0.35-1.20) |
| | vs BA.4/BA.5 vaccine | 46 (15.1) | 158 (20.3) | 0.94 (0.51-1.73) | 1.01 (0.50-2.03) |
| | vs wild-type vaccine | 98 (32.2) | 281 (36.0) | 0.78 (0.44-1.38) | 0.85 (0.43-1.66) |
| | vs unvaccinated | 113 (37.2) | 185 (23.7) | 0.45 (0.25-0.79) | 0.32 (0.16-0.63) |
| | XBB.1.5 vaccine ≥60 days earlier | 29 (9.5) | 90 (11.5) | — | — |
| | vs no XBB.1.5 vaccine | 257 (84.5) | 624 (80.0) | 0.78 (0.50-1.22) | 0.60 (0.35-1.04) |
| | vs BA.4/BA.5 vaccine | 46 (15.1) | 158 (20.3) | 1.11 (0.65-1.88) | 0.95 (0.50-1.79) |
| | vs wild-type vaccine | 98 (32.2) | 281 (36.0) | 0.92 (0.57-1.49) | 0.80 (0.44-1.46) |
| | vs unvaccinated | 113 (37.2) | 185 (23.7) | 0.53 (0.33-0.85) | 0.30 (0.17-0.55) |
| XBB (any) | XBB.1.5 vaccine 7-59 days earlier | 22 (2.2) | 126 (5.7) | — | — |
| | vs no XBB.1.5 vaccine | 956 (96.5) | 2,015 (91.5) | 0.37 (0.23-0.58) | 0.28 (0.17-0.47) |
| | vs BA.4/BA.5 vaccine | 166 (16.8) | 406 (18.4) | 0.43 (0.26-0.70) | 0.39 (0.23-0.68) |
| | vs wild-type vaccine | 429 (43.3) | 1,071 (48.6) | 0.44 (0.27-0.70) | 0.33 (0.19-0.56) |
| | vs unvaccinated | 361 (36.4) | 538 (24.4) | 0.26 (0.16-0.42) | 0.17 (0.10-0.29) |
| | XBB.1.5 vaccine ≥60 days earlier | 13 (1.3) | 61 (2.8) | — | — |
| | vs no XBB.1.5 vaccine | 956 (96.5) | 2,015 (91.5) | 0.45 (0.25-0.82) | 0.28 (0.14-0.55) |
| | vs BA.4/BA.5 vaccine | 166 (16.8) | 406 (18.4) | 0.52 (0.28-0.97) | 0.40 (0.19-0.81) |
| | vs wild-type vaccine | 429 (43.3) | 1,071 (48.6) | 0.53 (0.29-0.98) | 0.33 (0.16-0.67) |
| | vs unvaccinated | 361 (36.4) | 538 (24.4) | 0.32 (0.17-0.59) | 0.17 (0.08-0.34) |

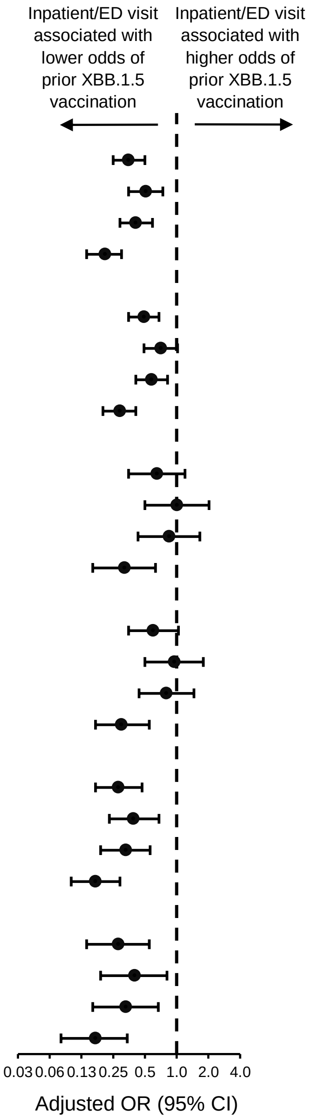


**Supplementary Figure 4. Association between inpatient or emergency department vs outpatient visit type and prior receipt of an XBB.1.5 vaccine 7-59 and ≥60 days earlier.** Associations were calculated among all SARS-CoV-2 infections and among JN.1 and XBB (any sublineage) infections. Odds ratios (ORs) were calculated comparing both receipt of an XBB.1.5 vaccine 7-59 days earlier and receipt of an XBB.1.5 vaccine ≥60 days earlier to no prior receipt of an XBB.1.5 vaccine (irrespective of previous COVID-19 vaccination history) as well as to each of three specific reference groups: 1) prior receipt of a BA.4/BA.5 vaccine but not an XBB.1.5 vaccine; 2) prior receipt of a wild-type vaccine but not a BA.4/BA.5 or XBB.1.5 vaccine; and 3) unvaccinated. Adjusted ORs were adjusted for age group (18-49, 50-64, 65-74, 75-84, and ≥85 years), sex, race and ethnicity (Asian, non-Hispanic; Black, non-Hispanic; Hispanic; white, non-Hispanic; and other/unknown), health system and state of residence, and collection date (natural cubic spline with 4 degrees of freedom). CI indicates confidence interval; ED, emergency department.

| SARS-CoV-2 variant | COVID-19 vaccination status | Inpatient and ED visits | Outpatient visits | OR (95% CI) | |
|---|---|---|---|---|---|
| | | No. (%) | No. (%) | Unadjusted | Adjusted |
| Any | Unvaccinated | 718 (37.6) | 1,124 (24.2) | 1.00 [Reference] | 1.00 [Reference] |
| | Wild-type vaccine | 783 (41.0) | 2,096 (45.2) | 0.58 (0.52-0.66) | 0.50 (0.43-0.59) |
| | BA.4/BA.5 vaccine | 294 (15.4) | 861 (18.6) | 0.53 (0.45-0.63) | 0.41 (0.33-0.50) |
| | XBB.1.5 vaccine | 117 (6.1) | 558 (12.0) | 0.33 (0.26-0.41) | 0.24 (0.19-0.32) |
| JN.1 | Unvaccinated | 113 (37.2) | 185 (23.7) | 1.00 [Reference] | 1.00 [Reference] |
| | Wild-type vaccine | 98 (32.2) | 281 (36.0) | 0.57 (0.41-0.79) | 0.38 (0.25-0.58) |
| | BA.4/BA.5 vaccine | 46 (15.1) | 158 (20.3) | 0.48 (0.32-0.71) | 0.32 (0.19-0.53) |
| | XBB.1.5 vaccine | 47 (15.5) | 156 (20.0) | 0.49 (0.33-0.74) | 0.31 (0.19-0.52) |
| XBB (any) | Unvaccinated | 361 (36.4) | 538 (24.4) | 1.00 [Reference] | 1.00 [Reference] |
| | Wild-type vaccine | 429 (43.3) | 1,071 (48.6) | 0.60 (0.50-0.71) | 0.51 (0.41-0.63) |
| | BA.4/BA.5 vaccine | 166 (16.8) | 406 (18.4) | 0.61 (0.49-0.76) | 0.42 (0.32-0.56) |
| | XBB.1.5 vaccine | 35 (3.5) | 187 (8.5) | 0.28 (0.19-0.41) | 0.17 (0.11-0.26) |
| HV.1 | Unvaccinated | 78 (33.9) | 135 (23.6) | 1.00 [Reference] | 1.00 [Reference] |
| | Wild-type vaccine | 97 (42.2) | 247 (43.1) | 0.68 (0.47-0.98) | 0.66 (0.42-1.04) |
| | BA.4/BA.5 vaccine | 46 (20.0) | 128 (22.3) | 0.62 (0.40-0.96) | 0.43 (0.24-0.74) |
| | XBB.1.5 vaccine | 9 (3.9) | 63 (11.0) | 0.25 (0.12-0.52) | 0.15 (0.06-0.34) |
| EG.5 | Unvaccinated | 85 (35.7) | 111 (21.9) | 1.00 [Reference] | 1.00 [Reference] |
| | Wild-type vaccine | 106 (44.5) | 282 (55.5) | 0.49 (0.34-0.70) | 0.56 (0.35-0.88) |
| | BA.4/BA.5 vaccine | 39 (16.4) | 81 (15.9) | 0.63 (0.39-1.01) | 0.49 (0.27-0.91) |
| | XBB.1.5 vaccine | 8 (3.4) | 34 (6.7) | 0.31 (0.14-0.70) | 0.22 (0.08-0.56) |

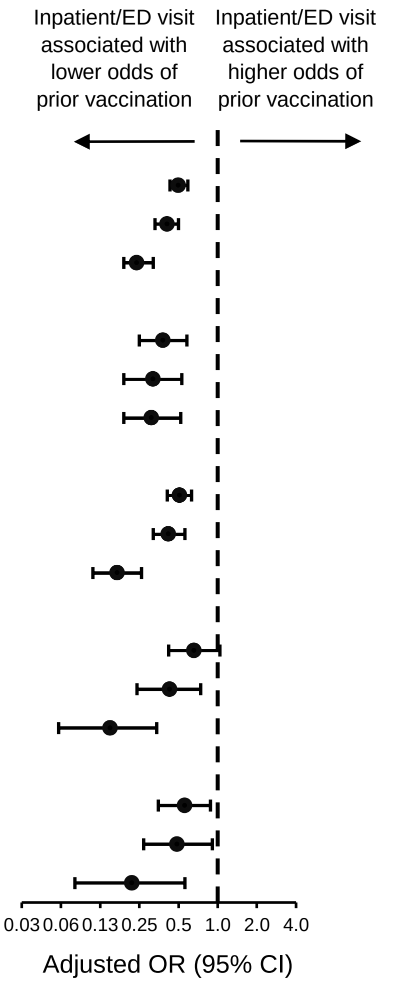


**Supplementary Figure 5. Association between inpatient or emergency department vs outpatient visit type and prior receipt of a wild-type, BA.4/BA.5, or XBB.1.5 vaccine, each vs unvaccinated.** Associations were calculated among all SARS-CoV-2 infections and among JN.1, XBB (any sublineage), HV.1, and EG.5 infections. Odds ratios (ORs) were calculated comparing each of three groups to unvaccinated: 1) prior receipt of a wild-type vaccine but not a BA.4/BA.5 or XBB.1.5 vaccine; 2) prior receipt of a BA.4/BA.5 vaccine but not an XBB.1.5 vaccine; and 3) prior receipt of an XBB.1.5 vaccine. Adjusted ORs were adjusted for age group (18-49, 50-64, 65-74, 75-84, and ≥85 years), sex, race and ethnicity (Asian, non-Hispanic; Black, non-Hispanic; Hispanic; white, non-Hispanic; and other/unknown), health system and state of residence, and collection date (natural cubic spline with 4 degrees of freedom). CI indicates confidence interval; ED, emergency department.
